# Deep Plasma Proteomics Reveals Shared and Disease-Specific Molecular Signatures in Alzheimer’s Disease and Frontotemporal Dementia

**DOI:** 10.64898/2026.04.14.26350728

**Authors:** Yi Jayne Tan, Monika Chauhan, Shourjo Chakravarty, Jigyasha Timsina, Muhammad Ali, Nicole Isabella Tan, Li Zeng, Louis CS Tan, Hui Jin Chiew, Kok Pin Ng, Shahul Hameed, Simon KS Ting, Jonathan D. Rohrer, Carlos Cruchaga, Bernett TK Lee, Adeline SL Ng

**Affiliations:** Department of Neurology, National Neuroscience Institute, Tan Tock Seng Hospital, 11 Jalan Tan Tock Seng 308433, Singapore; Lee Kong Chian School of Medicine, Nanyang Technological University, 11 Mandalay Road 30823, Singapore; Department of Psychiatry, Washington University School of Medicine, St. Louis, 63110, MO, USA; NeuroGenomics and Informatics Center. Washington University School of Medicine, St. Louis, 63110, MO, USA; Neural Stem Cell Research Lab, Department of Research, National Neuroscience Institute, Tan Tock Seng Hospital, 11 Jalan Tan Tock Seng 308433, Singapore; Department of Neurology, National Neuroscience Institute, Singapore General Hospital, Outram Road 169608, Singapore; Dementia Research Centre, Department of Neurodegenerative Disease, UCL Queen Square Institute of Neurology, University College London, Queen Square, London WC1N 3BG, United Kingdom; Neuroscience and Behavioural Disorders Programme, Duke-NUS Medical School, 8 College Road 169857, Singapore

**Keywords:** Biofluid, Biomarkers, Plasma, Alzheimer’s Disease, Frontotemporal Dementia, FTD, AD

## Abstract

**INTRODUCTION:** Alzheimer’s disease (AD) and frontotemporal dementia (FTD) have considerable clinical and pathological overlap. While plasma proteomics has advanced in AD, deep comparative analyses with FTD-particularly in diverse, biomarker-confirmed Asian cohorts-remain limited.

**METHODS:** Plasma from 101 individuals with known pTau217 status was profiled using Olink Explore-HT. Differential expression-pathway enrichment, penalized regression-GLMNET, single-cell transcriptomic integration, associations with cognitive measures and, cross-platform validation were performed.

**RESULTS:** Among 5,400-proteins, 1,168 were differentially expressed in AD and 370 in FTD (FDR<0.05). Distinct and overlapping proteomic signatures were identified in AD and FTD, reflecting gliosis, synaptic dysfunction, immune activation, and metabolic pathways. Prioritized proteins correlated with cognitive performance and plasma phosphorylated tau, Aβ_42_, and neurofilament light chain, linking circulating proteins to disease severity. Cross platform validation revealed strong concordance with large independent datasets.

**CONCLUSION:** Comprehensive plasma proteomics in Asian cohort supports scalable framework for blood-based biologically informed targets for precision diagnosis and therapeutic stratification.

## 1. BACKGROUND

Alzheimer’s disease (AD) and frontotemporal dementia (FTD) are the two most prevalent forms of dementia in younger-onset populations, characterized by overlapping clinical presentations but distinct neuropathological hallmarks. AD is primarily defined by the accumulation of amyloid-beta plaques and hyperphosphorylated tau tangles^1^, while FTD is pathologically and genetically heterogeneous, often associated with the aggregation of tau, TDP-43, or other proteins^2^, leading to the characteristic atrophy of the frontal and temporal lobes. The considerable clinical and pathological overlap between these diseases, particularly in their early stages, with atypical AD variants such as frontal variant AD that may resemble behavioural variant FTD, or overlapping features in logopenic primary progressive aphasia (PPA) typically associated with AD pathology vs semantic variant and non-fluent variant PPAs that typically associate with Frontotemporal Lobar Degeneration (FTLD) pathology, underscores the critical need for robust, non-invasive biomarkers to facilitate accurate differential diagnosis, patient stratification for clinical trials, and monitoring of disease progression. Despite distinct primary pathologies, both AD and FTD share fundamental underlying processes, including protein misfolding, aggregation, and progressive neuronal loss, which contribute to their shared clinical features.

Proteomics has emerged as a powerful tool for discovering novel biomarkers and uncovering the biological pathways that drive neurodegenerative diseases. While much of the early proteomic work focused on cerebrospinal fluid (CSF) due to its proximity to the central nervous system, recent advancements have enabled large-scale, high-throughput analysis of more accessible biofluids like plasma. To date, however, the majority of these proteomic efforts have focused specifically on characterizing AD. CSF proteomics have identified proteins related to protein catabolism, energy metabolism, and oxidative stress in AD^3^, whilst proteomic similarities between autosomal dominant and sporadic AD have been reported^4^, with a large-scale plasma proteomics study identified a high-performance biomarker panel for AD screening ^5^. More recently, high-throughput plasma proteomic studies have attempted to characterize and compare AD with FTD. A study using a limited panel of 1,303 proteins identified a 12-protein panel that accurately distinguished AD from FTD; however, this signature primarily reflected AD-associated changes rather than FTD-specific pathology^6^. In addition, a recent large-scale meta-analysis integrating aptamer-based proteomic datasets across multiple neurodegenerative disorders identified thousands of disease-associated proteins, revealing both shared and disease-specific molecular alterations^7^.

Despite these advances, comprehensive plasma proteomic studies comparing AD and FTD head-to-head are still limited, and the potential of a broad, unbiased analysis using the latest Olink platform, which assays over 5,000 proteins via antibody-based Proximity Extension Assay (PEA), remains unexplored. This is particularly crucial for providing cross-platform validation and ensuring that identified biomarkers are robust across different assay chemistries. In addition, the extent to which AD and FTD share common systemic molecular markers versus exhibiting disease-specific proteomics architecture is poorly defined. Only a few studies integrate plasma proteomics with brain derived disease specific single cell transcriptomics to infer cellular and mechanistic relevance of the circulating proteins. Compared to AD, FTD poses significant challenge in biomarker development as it includes heterogeneous groups of disorders characterized by tau, TDP-43 or FUS proteinopathies which are often associated with prominent synaptic loss and disruptions in nuclear and RNA-processing pathways. Whether these processes are captured in the plasma proteomics and how they differ from AD signatures is also an unexplored area.

In this study, we aim to bridge this gap in knowledge by conducting a large-scale, deep proteomic analysis of plasma from well-characterized AD and FTD patients using the Olink Explore HT platform (5,400 proteins). Our goal is to identify unique and shared protein profiles and their associated biological pathways to elucidate the distinct pathogenic mechanisms of each disease. By leveraging the latest high-throughput technology combined with pathway analysis, machine learning and cross platform validation, we seek to provide a foundation for developing a new generation of biologically informed blood-based biomarkers that can accurately differentiate AD from FTD, thereby improving diagnostic accuracy and accelerating the development of disease-modifying therapies.

## 2. METHODS

### 2.1 Clinical cohort

A total of 101 individuals with plasma measurements were included in the study: controls (n=50), FTD (n=11), AD (n=40). Patients were assessed and recruited from the tertiary memory clinics at the National Neuroscience Institute, Singapore between 2015 and 2024. This study was approved by the SingHealth Institutional Ethics Review Board and written informed consent was obtained from each participant prior to recruitment into the study. All patients underwent neurological examination, cognitive assessments, structural brain MRI, plasma pTau217 measurement and met consensus diagnostic criteria for AD dementia^8^ and FTD^9^ by neurologists specialized in dementia. All healthy controls (HCs) were free of significant neurological, psychiatric, or systemic disease. Patients with other neurodegenerative diseases such as Parkinson’s disease, with psychiatric comorbidities, as well as a history of alcohol or drug abuse were excluded. The Montreal Cognitive Assessment (MoCA^10^) and Mini Mental State Examination (MMSE^11^) were used to measure global cognitive function.

### 2.2 Plasma sampling and analysis

EDTA blood was centrifuged at 1,800 g for 10mins within 1 hour after collection. Plasma was aliquoted and stored at -80 °C until use. Plasma pTau217 was measured using ALZpath SIMOA p-tau217 Advantage PLUS kits (Quanterix, MA USA) on the HDX Analyzer, according to the manufacturer’s protocol. Established plasma pTau217 cutoffs were applied to support AD pathological classification^12,13^. AD participants had plasma pTau217 levels above the cutoff (>0.66 pg/ml), whereas FTD participants had pTau217 levels below this threshold.

### 2.3 Plasma proteomic profiling

Plasma samples were profiled using the Olink Explore HT platform, which applies antibody-based Proximity Extension Assay (PEA)^14^. The Explore HT panel enables multiplex quantification 5416 proteins in each sample and has demonstrated advantages in sensitivity, specificity and reproducibility compared to conventional antibody-based or mass spectrometry approaches. In PEA technology, paired antibodies are conjugated to unique oligonucleotides; when both antibodies bind in proximity to their target protein, the oligonucleotides hybridize and undergo proximity-dependent DNA ligation and elongation, generating a reporter sequence that is subsequently quantified by next-generation sequencing. Quality control and data normalization were performed using Olink’s standard pipeline, which includes sample control, inter-plate control, and normalization across all samples to minimize technical variation. Protein levels were converted into normalized protein expression (NPX) units, a relative quantification unit on a log_2_ scale. The normalized NPX were further normalized for the various batches by a median centering approach where the global and batch median of the sample control for each of the protein was computed. A batch offset was then computed which is the difference between the batch median to the global median. This batch offset was then applied to all samples within each of the batches to generate the finalized batch normalized data used for all subsequent analyses.

### 2.4 Statistical analysis

The statistical analysis of the data was conducted with the R programming language (version 4.5.1). T-tests between protein levels were conducted between HC, AD and FTD. False discovery rate (FDR) correction was conducted using the Benjamini-Hochberg method. Proteins passing a false discovery rate (FDR) threshold of less than 0.05 were considered significant. Subsequently, volcano plots and Venn diagrams based on the t-test were generated using the “ggplot” package for further visualisation.

In addition, the R “glmnet” package was used to produce a machine learning model for the prediction of AD and FTD based on differentially abundant proteins (DAPs) that were identified during the t-test (p<0.05). A binomial regression model with ten-fold cross validation was selected as the modelling approach. The end goal of producing these models was to highlight a set of proteins that work effectively together as a predictor for differentiating AD and FTD patients. Model coefficients were extracted to evaluate the relative contribution of each protein to classification.

A gene ontology (GO) enrichment analysis as well as a Reactome analysis were performed on the data sets as well. The gene ontology enrichment was carried out with the “clusterProfiler” package for R. The DAPs were split into two groups before the analysis: an AD group and an FTD group. The AD group contained all DAPs except for those unique to the FTD group – that is, the AD group contained the unique AD DAPs and the DAPs common to the FTD group and the AD group. Similarly, the FTD group consisted of the unique FTD DAPs along with the common DAPs. The GO analysis and Reactome analysis were then run individually on these two distinct groups. It is worth noting that we excluded DAPs without Entrez Identifiers at this stage because they were necessary for the packages used to run the GO and Reactome analysis. The common DAPs were included in both groups because it was found that without doing so, there would be insufficient DAPs to perform a meaningful analysis.

### 2.5 Cell typing analysis

Cell typing analysis for DAPs was done with the already published postmortem brain region specific single-cell transcriptomic datasets. The dataset for AD and FTD were accessed through the CZI CELLxGENE Discover platform^15^. Within the portal, AD/FTD related datasets were identified and filtered using the specific publications. For AD Entorhinal cortex dataset^16^ was used. For FTD dorsotemporal prefrontal cortex^17^ and frontal cortex^18^ datasets were used. The filtered dataset was selected on the platform and expression matrices corresponding to the top 30 DAPs and the GLMNET-selected proteins (for both AD and FTD) were extracted. CSV files exported from the CELLxGENE Discover gene-expression interface were imported into R for downstream processing. After loading required packages (“dplyr”, “readr”, “tidyr”, and “ggplot2”), each gene-cell type pair, summary statistics were computed, including the mean raw expression, mean scaled expression, total number of contributing cells, and the number of cells expressing each gene. These summaries were used to generate both a long-format results table and a gene by cell-type expression matrix which was used for visualization.

### 2.6 Cross-cohort and Platform Validation

For comparing our cohort to publish dataset from Ali et al 2025, the differential protein expression analysis was performed on R (4.5.1) to extract the effect size for each protein while adjusting for age and gender. Three pairwise contrasts were evaluated: 1. AD vs Control, 2. FTD vs Control and 3. AD vs FTD. Contrast matrices were constructed to estimate group differences for each comparison. Moderated t-statistics, raw p-values, and adjusted p-values were computed for all proteins. To account for multiple testing across the proteome, p-values were corrected using the Benjamini–Hochberg false discovery rate (FDR) procedure. Effect size results from our study and Ali et al. were imported and joined by performing an inner join on protein identifiers. For each matched protein, directionality of association (up- vs down-regulated in AD or FTD) was determined in both datasets, and concordance was defined as consistent directionality combined with statistical significance in both studies (false discovery rate [FDR] < 0.05). A filtered set of concordant proteins was derived for downstream cross-cohort validation analyses. Pairwise Spearman correlations between the AD effect sizes in our study and those reported by Ali et al. were also calculated to quantify agreement in effect sizes of both studies. The proteins were ranked based on the absolute effect-size differences (|β_Our − β_External|). These metrics were used to rank proteins, and the top concordant subset was visualized using bar plots and heatmaps. Scatterplots comparing effect sizes between cohorts were also generated with overlaid linear regression fits to highlight the global relationship between studies. Additional bar plots were created to highlight proteins with minimal cross-cohort effect-size divergence.

To assess the reproducibility of the proteomic signatures identified in our Olink Explore HT dataset, another cross-platform comparison was performed with independent Somascan 7K proteomic datasets from plasma (N=733) and cerebrospinal fluid (CSF; N=2,183) from the Charles F. and Joanne Knight Alzheimer Disease Research Center (Knight ADRC) Cohort housed at Washington University in St. Louis^19,20^. Processed differential abundance results from Somascan 7k assays were obtained for plasma and CSF samples using same models, including comparisons of AD vs HC, FTD vs HC, and AD vs FTD. All Somascan datasets had undergone internal quality control as described previously^19,20^. Since, Somascan platform includes multiple aptamers for some proteins, analyses were conducted at both analyte and protein levels. First, Olink proteins were mapped to Soma targets using entrez gene and UniProt symbols and the Somascan DAPs list was created. If multiple Somascan analytes corresponded to a single protein, each analyte was retained and tested independently. Then, significant proteins from our cohort were intersected (using Venn diagrams) with the list of proteins measurable in the Somascan plasma and CSF panels. This defined the total testable overlap.

## 3. RESULTS

### 3.1 Participant characteristics

Demographic information and clinical characteristics of the total participants are presented in Table 1. Plasma phosphorylated tau levels (pTau217 and pTau181) were significantly higher in AD as compared to both FTD and controls, consistent with the established AD-specific pathology^13^.

### 3.2 Distinct and overlapping proteomic changes in AD and FTD

Among the 5,400 assayed plasma proteins, 1,168 were differentially altered in patients with AD, including 622 significantly upregulated proteins and 546 significantly downregulated proteins (FDR-adjusted p<0.05; Figure 1A and Supplementary Table S1 in supporting information). In patients with FTD, 370 proteins were differentially expressed (FDR-adjusted p<0.05; Figure 1B and Supplementary Table S2), of which 170 are significantly upregulated and 200 are significantly downregulated.

**Figure 1.**
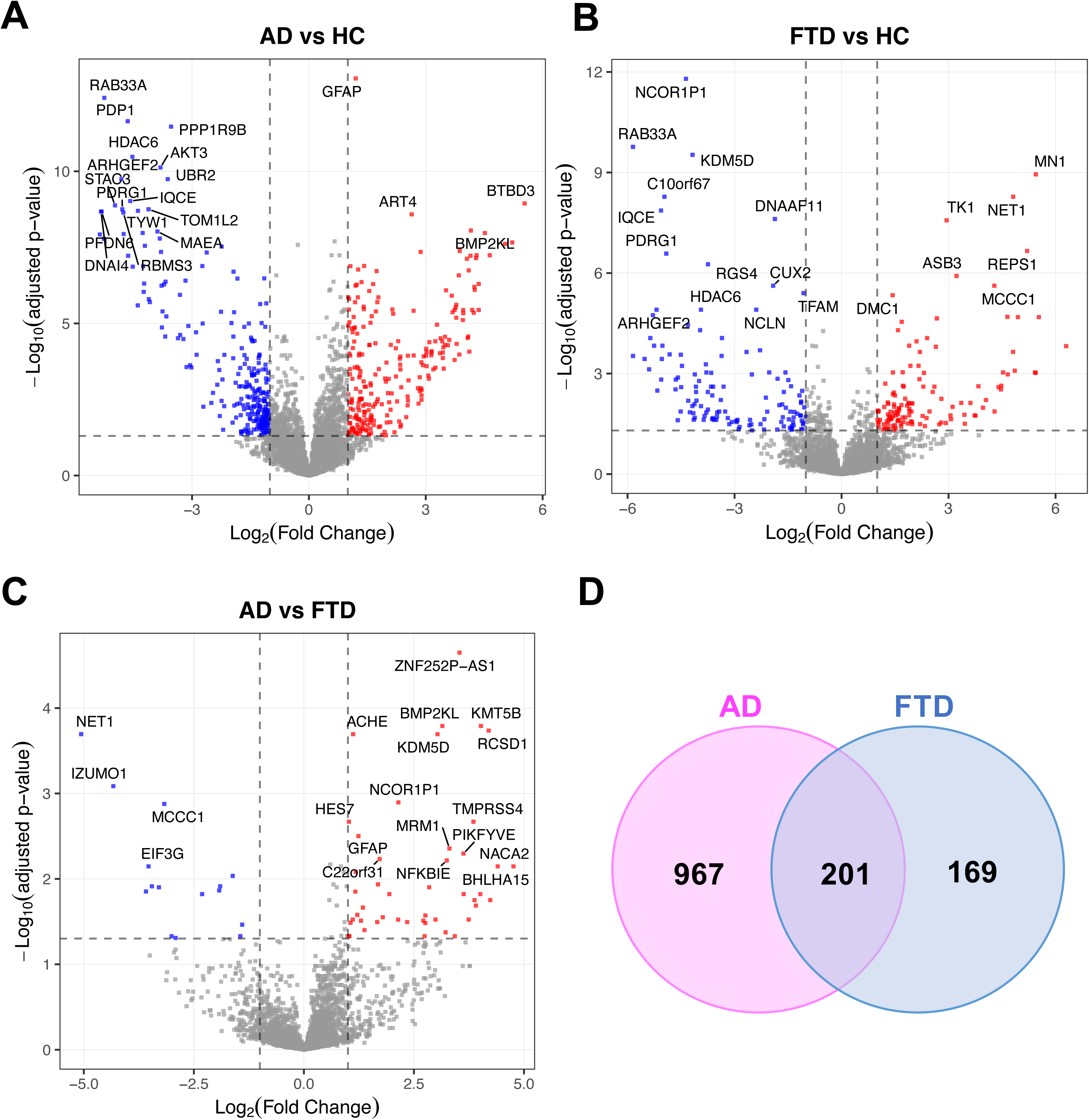
Differential plasma proteomic signatures distinguishing AD and FTD. Volcano plots of differentially abundant proteins (DAPs) in (A) AD vs Control, (B) FTD vs Control and (C) AD vs FTD; (D) Venn diagram showing overlap od DAPs in AD and FTD relative to HC. The x-axis represents log□ fold change, and the y-axis shows −log□□ (adjusted p-value). Red dots indicate significantly upregulated proteins, blue dots indicate significantly downregulated proteins, and grey dots represent non-significant proteins. Selected top-ranking proteins are annotated. Among 5400 plasma proteins assayed, 1168 and 370 were differentially expressed in AD and FTD as compared to HC, respectively. Of these, 201 proteins (15%) were shared between the two disorders, with ∼48% consistently increased, 52% consistently decreased, and 0.5% showing opposite directionality of change.

Notably, AD exhibited larger number of disease-specific proteins (967, 72% of all DAPs), compared to FTD (169, 13%). A total of 201 proteins (15%) were shared between AD and FTD. Among those shared proteins, 96 (48%) were consistently increased; 104 (52%) were consistently decreased; and 1 (0.5%) showed opposite directionality of change.

#### 3.2.1 Top Differentially Altered Proteins in AD and FTD

Among the top differentially regulated proteins in AD, there are several established markers and disease relevant proteins (Figure 1C), including GFAP (astrocyte marker, *q* = 8.8×10^-14^)^21^, BACE1 (central β-secretase in Aβ generation, *q* = 2.1×10^-8^)^22^, PPP1R9B (dendritic-spine protein, *q* = 3.4×10^-12^)^23^, HDAC6 (histone deacetylase implicated in tau processing, q = 3.3×10^-11^)^24^, BECN1 (autophagy protein, *q* = 2.0×10^-7^)^25^, SYT1 (pre-synaptic protein, q = 7.7×10^-7^)^26^, and ACHE (acetylcholinesterase, *q* = 1.2×10^-5^). In FTD, differentially regulated proteins included NEFL (axonal injury marker, *q* = 9.18×10^-4^), HDAC6 (*q* = 1.25×10^-5^), TFAM (mitochondrial stress, q = 4.0×10^-6^), EIF3G (translation initiation, *q* = 2.0×10^-5^), the RNA/DNA processing proteins DDX53 (q = 2.1×10^-5^) and KDM5D (q = 3.0×10^-10^).

#### 3.2.2 Pathway Enrichment Analysis of AD-Specific Proteins

To explore the biological processes underlying proteins uniquely altered in AD, we performed enrichment analyses using Gene Ontology (GO) Biological Process (BP) and Reactome databases.

GO BP analysis of the highlighted significant enrichment of pathways related to mitochondrial dysfunction (mitochondrial respiratory chain complex assembly, NADH dehydrogenase complex assembly), programmed necrotic cell death, and microtubule organization and polymerization. Several terms involved in synaptic and neurotransmitter processes (synaptic vesicle exocytosis, neurotransmitter secretion) and amyloid-related processes (amyloid precursor protein metabolic process, amyloid-beta formation) were also overrepresented (Figure 2A, Supplementary Table S3.1). These findings suggest that mitochondrial impairment, regulated necrosis, cytoskeletal instability, and synaptic dysfunction converge in AD-specific pathology.

**Figure 2.**
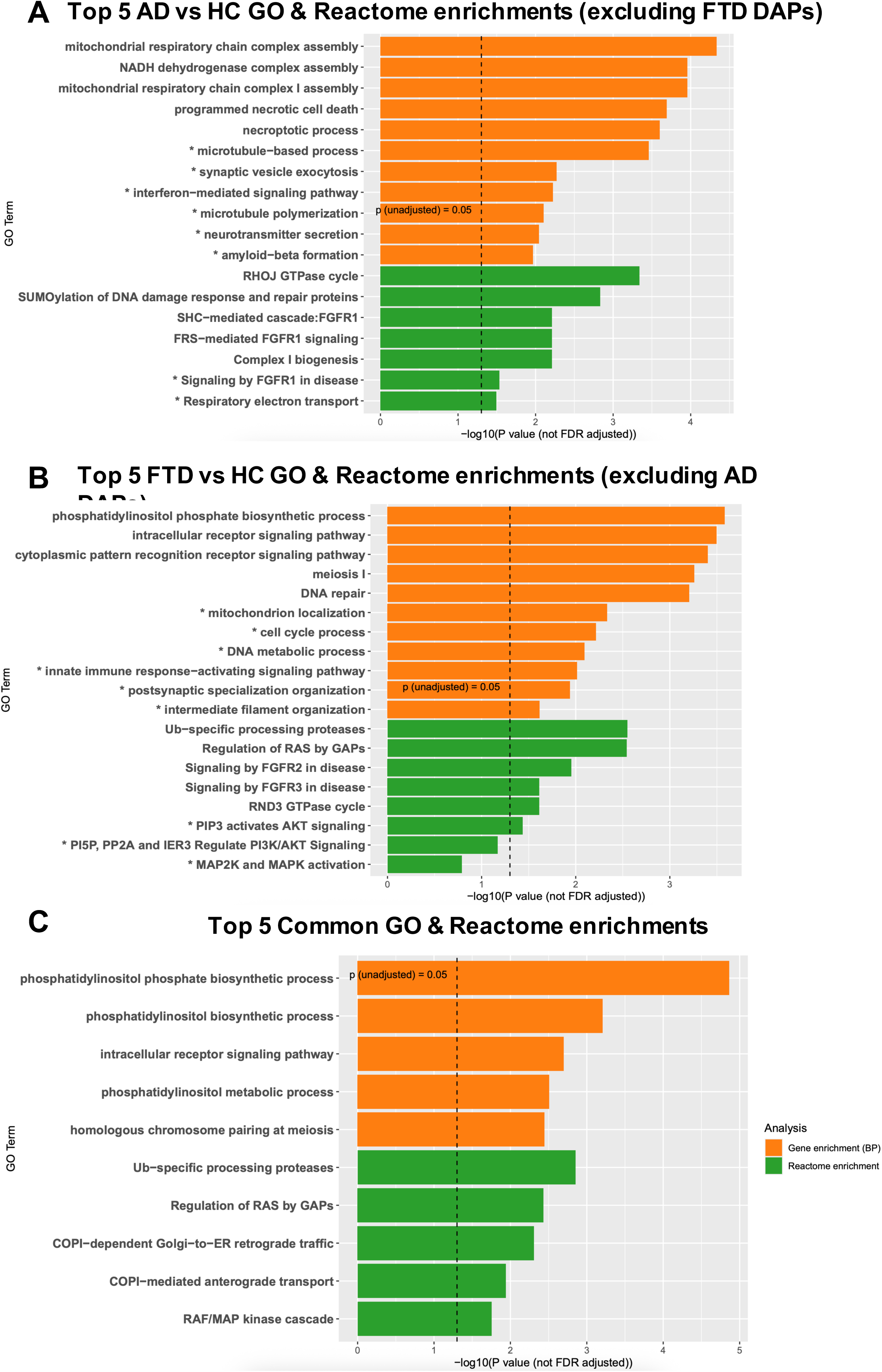
Biological Pathway analysis of DAPs in AD and FTD. (A) Significant AD vs HC DAPs excluding the common ones significant in FTD vs HC were subjected to pathway enrichments. (B) Significant FTD vs HC DAPs excluding the ones significant in AD vs HC were subjected to pathway enrichment. (C) The common DAPs in both lists (AD vs HC and FTD vs HC) were subjected to pathway enrichments. Top 5 significant GO and Reactome pathways are presented along with selected disease specific pathways (marked with *).

Reactome pathway analysis corroborated these results and provided further resolution. Top enriched pathways included RHO GTPase signalling (RHOJ and RHOQ GTPase cycles), Complex I biogenesis, and respiratory electron transport, reinforcing mitochondrial and cytoskeletal abnormalities. Enrichment of FGFR1 signalling (FRS- and SHC-mediated cascades) and SUMOylation of DNA repair and RNA-binding proteins pointed to dysregulated growth factor signalling and post-translational modifications as additional contributors to AD pathology (Figure 2A, Supplementary Table S3.1).

Together, these analyses indicate that proteins uniquely dysregulated in AD are involved in a coordinated network of mitochondrial dysfunction, cytoskeletal disorganization, regulated cell death, synaptic impairment, and aberrant growth factor/post-translational signalling, consistent with core mechanisms of AD pathogenesis.

#### 3.2.3 Pathway Enrichment Analysis of FTD-Specific Proteins

Pathway enrichment of proteins uniquely dysregulated in FTD compared with healthy controls revealed distinct biological signatures compared with AD.

GO Biological Process analysis highlighted enrichment of DNA metabolic and repair processes, cell cycle regulation, and meiosis-related pathways, pointing to alterations in nuclear integrity and genomic stability. In addition, innate immune response–activating signalling, cytoplasmic pattern recognition receptor signalling, and postsynaptic specialization organization were significantly enriched, implicating immune dysregulation and synaptic structural changes in FTD (Figure 2B, Supplementary Table S3.2).

Reactome pathway analysis further revealed strong enrichment of RAS–RAF–MAPK and PI3K/AKT signalling cascades^7,27,28^, including signalling by BRAF and RAF1 fusions, signalling by RAF1 mutants, MAP2K and MAPK activation, and PI3K/AKT signalling regulation. FGFR2-mediated signalling pathways (FRS-mediated FGFR2, SHC-mediated FGFR2, Signalling by FGFR2 in disease) were also enriched, suggesting aberrant growth factor signalling (Figure 2B, Supplementary Table S3.2). Finally, pathways linked to ubiquitin-specific processing proteases were overrepresented, consistent with dysregulation of protein homeostasis^7,27,28^.

Together, these findings indicate that FTD-unique proteins converge on DNA/nuclear processes, immune signalling, and aberrant growth factor and kinase signalling, distinguishing the molecular underpinnings of FTD from those of AD. Taken together, these results suggest that proteins uniquely dysregulated in FTD are linked to DNA and nuclear functions, innate immune pathways, cytoskeletal remodelling, and postsynaptic structure, reflecting disease mechanisms distinct from those identified in AD.

#### 3.2.4 Pathway analysis of overlapping proteins in AD and FTD

The pathway enrichment of proteins shared between AD and FTD revealed a focused set of biological processes centred on phosphoinositide metabolism, vesicle trafficking, and protein quality control, highlighting mechanisms common to both neurodegenerative disorders. GO terms such as phosphatidylinositol biosynthetic and metabolic processes point to shared disruption of lipid signalling pathways that govern membrane identity, autophagy, and synaptic vesicle dynamics, while Reactome pathways including COPI-mediated anterograde and retrograde transport and regulation of RAS by GAPs indicate impairments in Golgi–ER trafficking and small GTPase signalling. Enrichment of ubiquitin-specific processing proteases and the RAF/MAP kinase cascade further suggests overlapping disturbances in proteostasis and stress-activated kinase pathways (Figure 2C, Supplementary Table S3.3). Together, these findings show that despite disease-specific pathways in AD and FTD, the overlapping proteins converge on fundamental cellular processes involving lipid signalling, intracellular transport, and ubiquitin-mediated protein regulation—representing shared molecular vulnerabilities in neurodegeneration.

### 3.3 GLMNET identifies discriminatory protein panels for AD and FTD

Using a GLMNET penalized regression model, we identified a panel of proteins that best distinguished AD from HC (Figure 3A, Supplementary Table S4.1). The model retained both positively and negatively associated proteins, with the direction and magnitude of coefficients reflecting their contribution to classification. Among the strongest positive predictors were GFAP (2.03), BACE1 (0.85), N4BP2L2 (0.49), ACHE (0.48), and APOA2 (0.25), which were elevated in AD compared to controls. Conversely, proteins such as FLT3LG (-1.02), PPP1R9 (-0.36), and VGF (-0.35) showed negative coefficients, indicating relatively lower levels in AD. GFAP emerged as the most influential feature with the highest positive coefficient, while FLT3LG was the strongest negative predictor (Figure 3A). These results highlight a subset of proteins with potential discriminative value for AD classification from proteomic profiling. Additionally, GLMNET also selected several proteins with smaller coefficients. While individually these proteins may contribute less to classification, their selection indicates potential additive value within the multivariate model. Many are involved in pathways broadly relevant to AD, such as cytoskeletal regulation^29,30^ and neuroinflammation^31^ (e.g., CCNE1, TNFSF10, SPTBN4). Others are less characterized in neurodegeneration, representing possible novel candidates that warrant further investigation. The GLMNET model for FTD vs HC identified a panel of proteins reflecting neuroinflammation, axonal injury, and cellular stress in FTD (Figure 3B, Supplementary Table S5.1). The strongest contributors, C3 (0.85) and NEFL (0.70), correspond to complement activation^32^ and neurofilament release^33^ well-established markers of inflammation and axonal degeneration. Other retained proteins, including LTA4H (0.08), AKT3 (-0.08), TFAM (-0.18), and BCL2L11 (-0.35), implicate lipid-mediated inflammation^34^, mitochondrial dysfunction^35,36^, and apoptosis^37^. Additional features such as PPFIA2 (-0.03), NTF4 (-0.02), and C10orf67 (-0.05) suggest contributions from synaptic remodelling^38^, neurotrophic support^39^, and hypoxic stress^40^, indicating that a combination of diverse molecular pathways optimally distinguishes FTD from controls.

**Figure 3.**
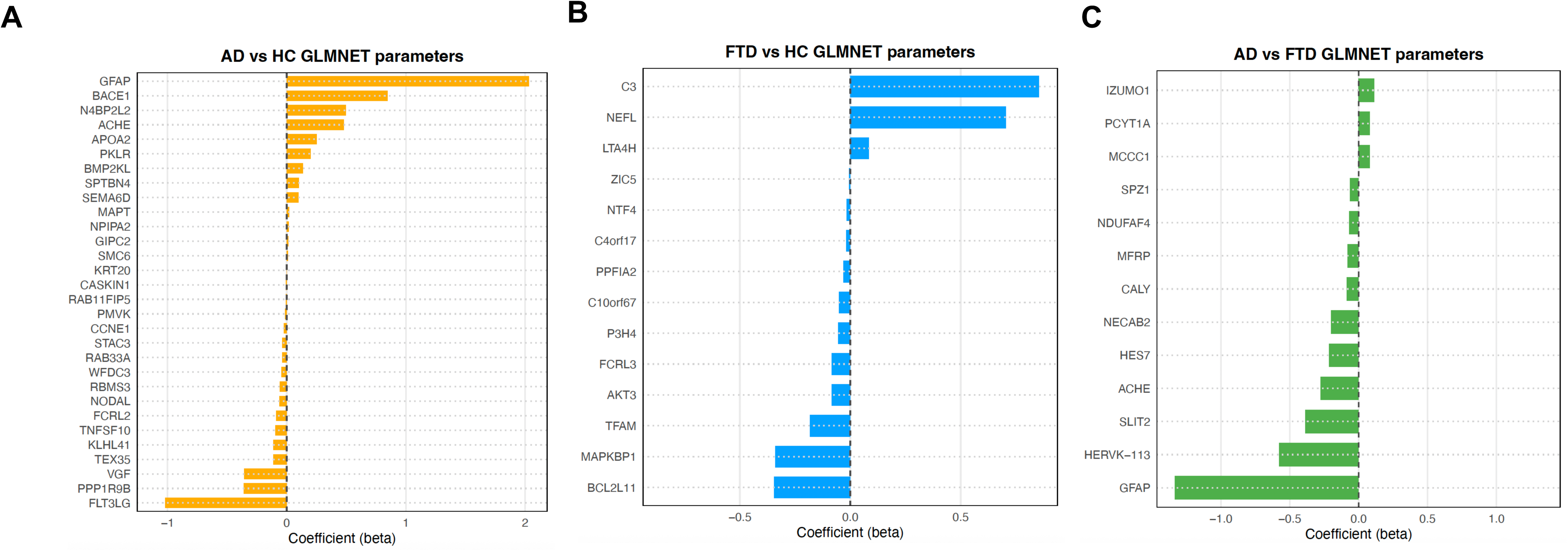
GLMNET-selected proteins in AD and FTD - Individual protein coefficients. GLMNET-derived multivariate model was applied to the dataset to identify DAPs contributing to the disease. Coefficients for proteins selected by GLMNET for distinguishing (A) AD from HC, (B) FTD from HC, and (C) AD from FTD. Positive coefficients indicate proteins elevated in AD or FTD relative to controls, whereas negative coefficients indicate proteins reduced in AD or FTD. The magnitude of the coefficient reflects the relative contribution of each protein to the classification model. Only proteins retained after regularization are presented.

The AD vs FTD GLMNET model revealed a distinct set of discriminatory proteins, largely driven by markers elevated in AD (Figure 3C, Supplementary Table S6.1). GFAP (-1.33) and ACHE (-0.27), both retained in the AD vs controls model, were again among the strongest features, highlighting consistent astroglial activation and cholinergic pathway involvement as AD-specific signals. Several proteins, including SLIT2 (-0.38), NECAB2 (-0.19), HERVK–113 (-0.58), CALY (-0.08), MFRP (-0.07), and NDUFA4 (-0.07), were uniquely selected in the AD vs FTD model, suggesting potential disease-specific differences in synaptic signalling, mitochondrial metabolism, and astrocytic regulation. These newly identified proteins may represent novel molecular discriminators between AD and FTD and warrant further mechanistic investigation.

### 3.4 Associations between clinical/biomarker measures and key AD and FTD plasma proteins

To evaluate the clinical relevance of the prioritized proteomic signature, we examined the associations between the top 30 DAPs and GLMNET-selected features with key cognitive and biochemical markers in AD and FTD. A substantial proportion of these proteins demonstrated significant associations with cognitive performance and plasma AD biomarkers (Figure 4).

**Figure 4.**
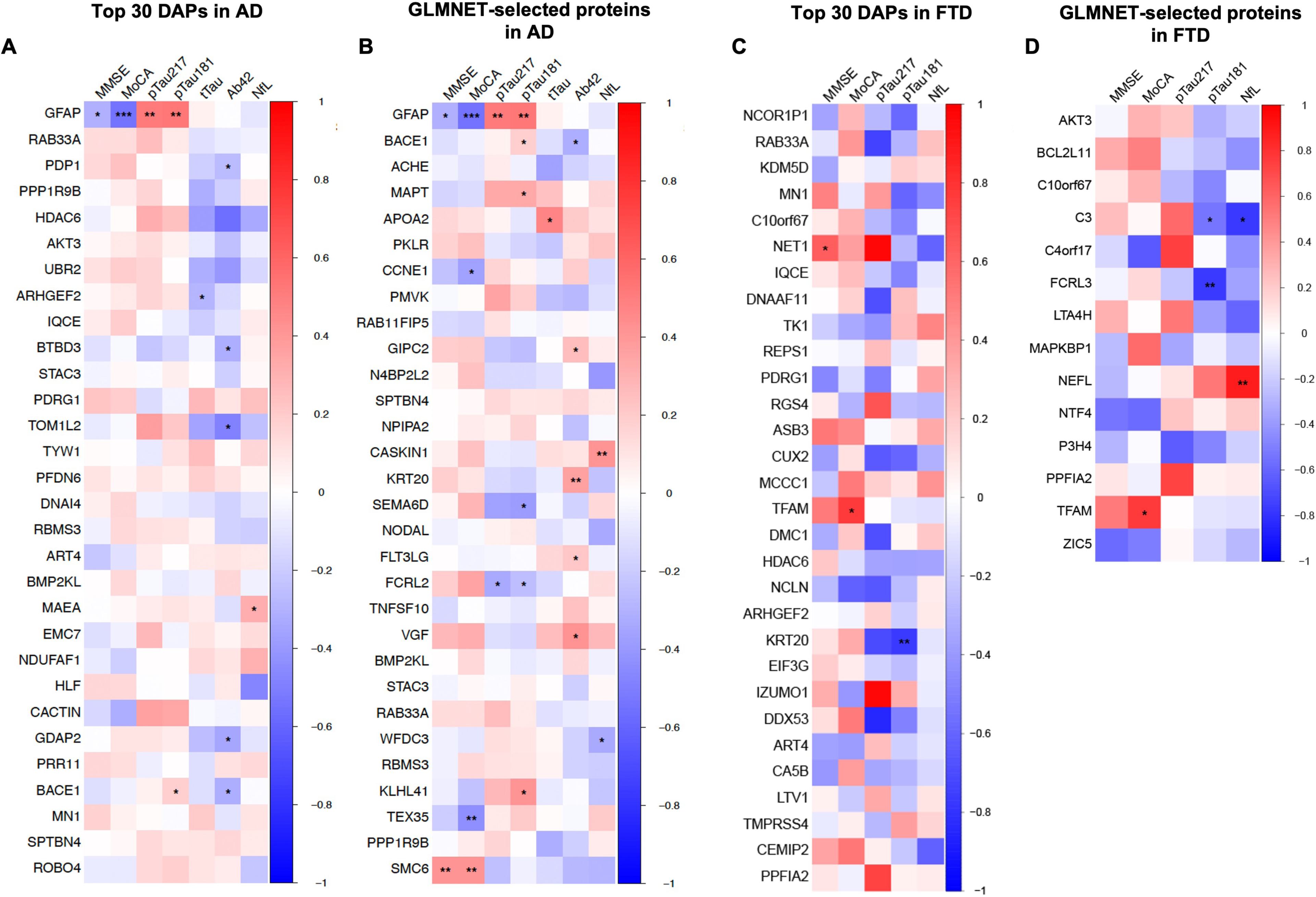
Correlation of associated plasma proteins with clinical and biochemical variables. Heatmaps of Spearman correlation coefficients between (A) the top 30 differentially abundant proteins (DAPs) in AD, (B) GLMNET-selected proteins in AD, (C) top 30 DAPs in FTD, (D) GLMNET-selected proteins in FTD and clinical/biomarker variables, including MMSE, MoCA, plasma pTau217, pTau181, total tau (tTau), Aβ42, and neurofilament light (NfL). The color gradient represents the strength and direction of the correlation (Red = positive; Blue = negative), ranging from −1 to +1. Asterisks denote statistically significant correlations (*** p<0.001, **p<0.01 and *p<0.05)

Among the top-ranked proteins in AD, GFAP showed significant associations with both cognitive measures (MMSE and MoCA) and phosphorylated tau species (pTau217 and pTau181). Within the GLMNET-selected panel, several proteins exhibited relationships with cognitive scores, including CCNE1, TEX35, and SMC6. Regarding biochemical markers of pathology, BACE1, MAPT, KLHL41, SEMA6D, and FCRL2 demonstrated associations with pTau181 or pTau217. Total tau (tTau) levels were linked to APOA2, ARHGEF2, and GDAP2. Furthermore, a subset of proteins, including PDP1, BTBD3, TOM1L2, GIPC2, KRT20, FLT3LG, and VGF, showed significant associations with Aβ_42_. Finally, relationships with NfL were observed for MAEA, CASKIN1, and WFDC3, highlighting proteins potentially linked to neuroaxonal injury.

Among the top-ranked proteins in FTD, NET1 demonstrated a significant association with MMSE scores. KRT20 was significantly associated with pTau181 levels. Within the GLMNET–selected panel, significant associations with pTau181 were observed for C3 and FCRL3. Furthermore, C3 exhibited a significant association with NfL, highlighting its relationship with established markers of neurodegeneration in the FTD cohort.

Collectively, this demonstrates that proteins selected through differential abundance and GLMNET are not only disease associated but also track the cognitive impairment and core pathology indicating their potential use as clinically informative biomarkers.

### 3.5 Cell-Type Specificity Using Single-Nucleus Transcriptomics

Top 30 proteins and GLMNET selected proteins were integrated with the datasets of single-cell transcriptomic data from AD post-mortem human entorhinal cortex brain samples with varying degrees of AD pathology and cognitive impairment^15,16^. Top 30 proteins and GLMNET selected proteins in FTD dataset were integrated with the datasets of single-cell transcriptomic data from FTD post-mortem human frontal cortex^18^ and dorsolateral prefrontal cortex^17^ brain samples with varying degrees of FTD pathology^15^.

To determine the potential cellular origins of AD–associated plasma proteins, we examined single-cell RNA-sequencing data from the entorhinal cortex region of AD and control post-mortem brain tissues^15,16^ focusing on top 30 DAPs and proteins selected by GLMNET analysis. The analysis revealed distinct and cell type–specific expression patterns across neuronal, glial, and vascular populations (Figure 5).

**Figure 5.**
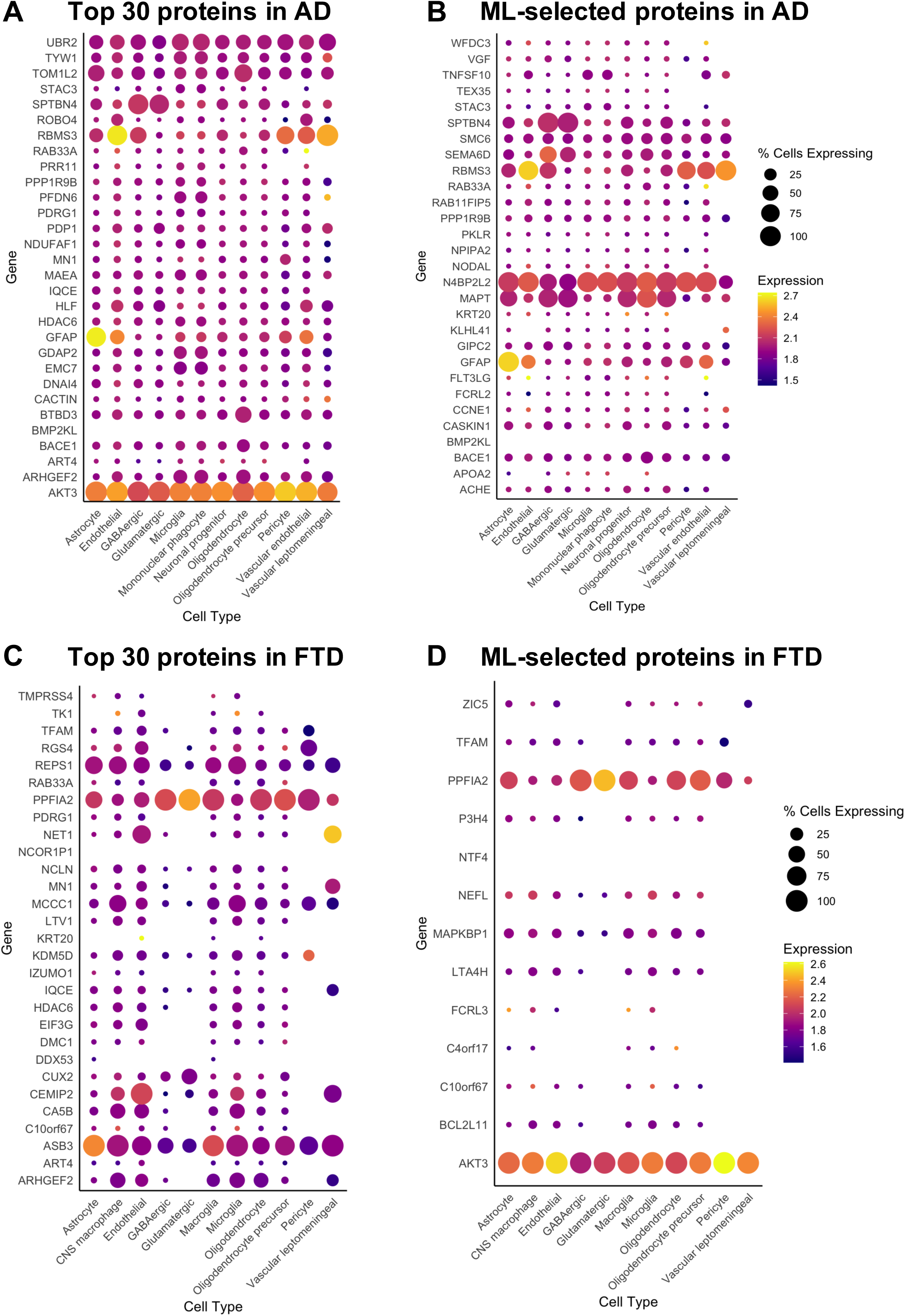
Cellular origin and mapping of plasma proteomic signatures using single-cell transcriptomic integration. Dot plots indicating cellular origins of disease-associated plasma proteins by integration with scRNA-sequencing datasets derived from post-mortem disease specific human brain tissue. Each panel corresponds to a disease comparison: (A) AD vs HC Top 30 DAPs; (B) AD vs HC GLMNET-selected proteins. Top 30 DAPs and GLMNET selected proteins were integrated with the datasets of single-cell transcriptomic data from AD entorhinal cortex brain samples with varying degrees of AD pathology and cognitive impairment. (C) FTD vs HC Top 30 proteins; (D) FTD vs HC GLMNET-selected proteins. Top 30 proteins and GLMNET selected proteins in FTD dataset were integrated with the datasets of single-cell transcriptomic data from FTD post-mortem human frontal cortex and dorsolateral prefrontal cortex brain samples with varying degrees of FTD pathology. Genes (y-axis) represent selected DAPs identified in plasma, while cell types (x-axis) include major cell types. Dot size reflects the percentage of cells expressing the gene within each cell type, and color intensity represents normalized expression levels.

Most proteins, such as AKT3 and N4BP2L2, exhibited widespread expression across all cell-types, suggesting a role in fundamental cellular processes or systemic pathological involvement. However, the analysis also captured highly specialized expression profiles. In the neuronal compartment, SPTBN4 and MAPT were distinctly enriched in GABAergic and glutamatergic neurons, while SEMA6D showed elevated expression specifically within the GABAergic population. GFAP was exclusively enriched in astrocytes, BTBD3 showed restricted expression within the oligodendrocyte lineage, and RBMS3 served as a distinct marker for vascular leptomeningeal cells and pericytes. Additionally, ARHGEF2 displayed a preferential expression pattern in glial populations, with higher prevalence observed in microglia and oligodendrocytes compared to neurons. Conversely, BMP2KL showed minimal expression across all surveyed brain cell-types (Figure 5A, 5B, Supplementary Table S7.1, S7.2).

These findings suggest that plasma proteins differentiating AD from controls are derived from diverse brain cell populations in the entorhinal cortex, reflecting contributions from neuronal, glial, and vascular compartments that collectively mirror the complex cellular pathology of AD.

A similar integrative analysis was carried out for FTD specific 30 DAPs and proteins selected by GLMNET with single-cell RNA-sequencing datasets from FTD post-mortem frontal cortex and dorsolateral prefrontal cortex tissues^17,18^. ASB3 expression was highest in glial populations, AKT3 was expressed highest in the endothelial and pericytes while PPFIA2 were highly expressed in glutamatergic neurons. AKT3 and PPFIA2 were also enriched in broader cell types. FCRL3, MCCC1, C10rf67, C4orf17 showed selectively higher expression in the immune cells indicating a possible contribution of innate immune cells to FTD associated plasma proteome^41,42^ (Figure 5C, 5D, Supplementary Table S7.3, S7.4).

### 3.6 Cross-platform and external validation of the plasma proteome

#### 3.6.1 Validation in the GNPC SomaScan cohort

To evaluate the robustness of our results, we compared our Olink-derived effect sizes (β) with a large published SomaScan 7k dataset from Ali et al. (2025)^7^, generated from 1,936 AD cases and 6,265 controls within the GNPC cohort.

This cross-platform validation for AD demonstrated strong concordance, with 232 (19.86% of total significant) proteins showing consistent directional changes (FDR<0.05) across both cohorts (Figure 6A). Effect sizes were strongly correlated (Spearman ρ=0.76; Supplementary Figure 1A), indicating that the disease-associated proteomic signal is stable and assay-independent. Furthermore, ranking proteins by the absolute difference in effect size (Delta Effect size) identified markers with exceptional reproducibility, including HK2, TNFRSF14, ENG, and PECAM1 (Figure 6B, Supplementary Table S8.1).

**Figure 6.**
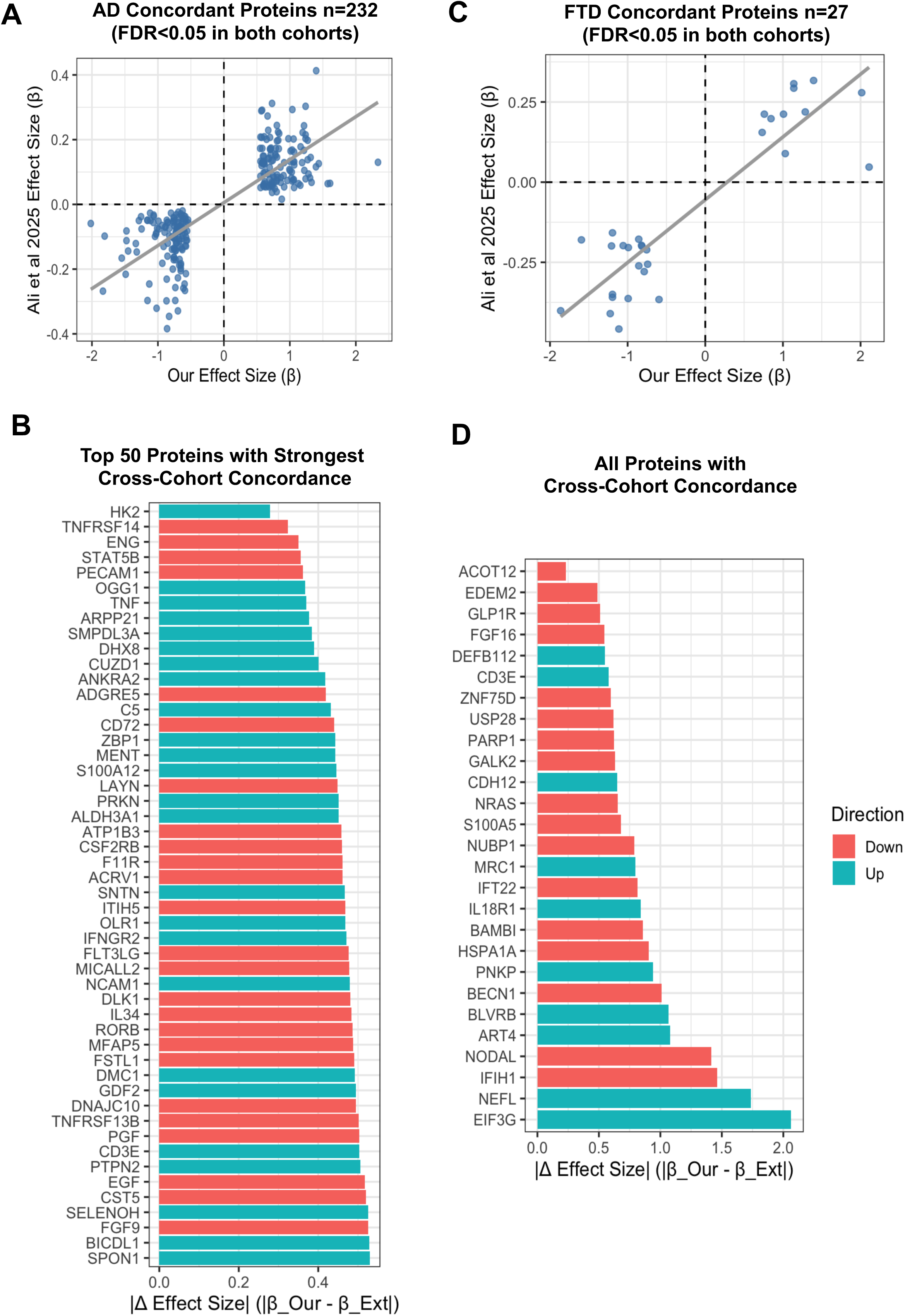
Cross-cohort and platform validation. FDR <0.05 selected DAPs in our dataset were compared with external published data. Scatter plot comparing log□ fold-change effect sizes (β) from the present study (“Our”) with those reported in an independent external cohort^7^ for (A) AD vs HC and (B) FTD vs HC. Each point represents a shared protein between platforms. Gray regression line indicates the linear relationship between studies. Dashed lines denote zero effect in each dataset. A strong positive association demonstrates concordant direction and magnitude of protein alterations across cohorts. Bar plot of top 50 shared proteins in AD (C) and FTD (D) ranked by absolute effect size difference (|β_Our − β_Ext|). Bars are colored according to direction of change (Up = teal; Down = red). The majority of proteins show consistent directionality, with only slight effect size divergence across cohorts.

Pathway enrichment analysis of the 232 concordant proteins revealed reproducible biological signature of AD (Supplementary Figure 1B). The most significant enrichments were found in immune and inflammatory signalling (cytokine signalling, TNF-mediated signalling), cell death and survival (regulation of necroptotic process, cell population proliferation), and growth factor and receptor kinase activity (FGFR1 signaling, tyrosine kinase binding), with secondary involvement of vascular and cell structure pathways. This shared signature suggests that the stable pathology captured in plasma is centered on chronic neuroinflammation and dysregulated cellular viability (Supplementary Figure 1B).

Analysis of protein directionality showed a clear dichotomy (Supplementary Table S8.1), reflecting the simultaneous occurrence of injury and homeostatic failure. The upregulated cluster was dominated by markers of metabolic stress (HK2), neuroinflammation (S100A12, TNF), and genomic instability (DHX8, OGG1), while the downregulated cluster primarily consisted of markers associated with vascular integrity (PECAM1, ENG), immune regulation (TNFRSF14). This segregation confirms that the reproducible AD plasma profile is defined by the elevation of stress and injury markers and the concurrent suppression of essential protective and homeostatic mechanisms.

FTD dataset also demonstrated strong concordance, with 27 (7.29% of total significant proteins) proteins showing consistent directional changes (FDR<0.05) (Figure 6C, Supplementary Table S8.2). Effect sizes were strongly correlated between cohorts (Spearman ρ=0.74; Supplementary Figure 2A), indicating that the disease-associated proteomic signal is stable and assay-independent for FTD as well. Several proteins, including ACOT12, EDEM2, and GLP1R, showed minimal absolute differences in effect size, further supporting robust cross-platform reproducibility (Figure 6D). Pathway enrichment of the concordant proteins revealed significant involvement of FGFR2/3/4 and IGF1R signaling, ubiquitin-processing pathways, DNA damage repair, and transition metal ion binding (Supplementary Figure 2B). Together, these cross-validated proteins delineate a reproducible molecular architecture of FTD, centred on axonal degeneration (NEFL), impaired stress-response and DNA repair (EIF3G, PARP1, USP28), altered growth-factor signaling (FGF16, GLP1R), and metabolic dysregulation (ACOT12, BLVRB). This convergence across independent datasets confirms that a biologically coherent FTD signature is detectable in plasma and remains consistent across proteomic platforms.

#### 3.6.2. Validation in the Knight ADRC Plasma and CSF Cohorts

We then compared our Olink Explore HT plasma with another independent external dataset using SomaScan 7k on plasma and CSF (Figure 7). In plasma-CSF comparison, among 1,168 FDR-significant AD proteins in our Olink plasma dataset, 509 unique proteins (599 Somascan analytes) were present in Somascan CSF, of which 374 analytes passed the FDR correction in the CSF dataset. Among these, 21 proteins overlapped across all three (Olink plasma, Somascan plasma and Somascan CSF) datasets in AD vs HC, several have documented associations with AD -SYT1^26^, SPP1^43^, APOE^44^, ACHE^45^, NEFL, TFRC^46^, SPON1^47^, SERPINA1^48^ and IGF1R^49^- supporting biological relevance and cross-platform reproducibility of our findings. In plasma-plasma comparison, 502 unique proteins were analysed, with 30 Somascan analytes reaching FDR significance, indicating that a subset of AD associated proteins can be reproducibly detected across different assay chemistry technologies and target design. 314 proteins overlapped across Olink 5k plasma and Somascan 7k plasma datasets. The remaining overlapped proteins are largely understudied in AD and represent promising novel candidates for validation (Figure 7).

**Figure 7.**
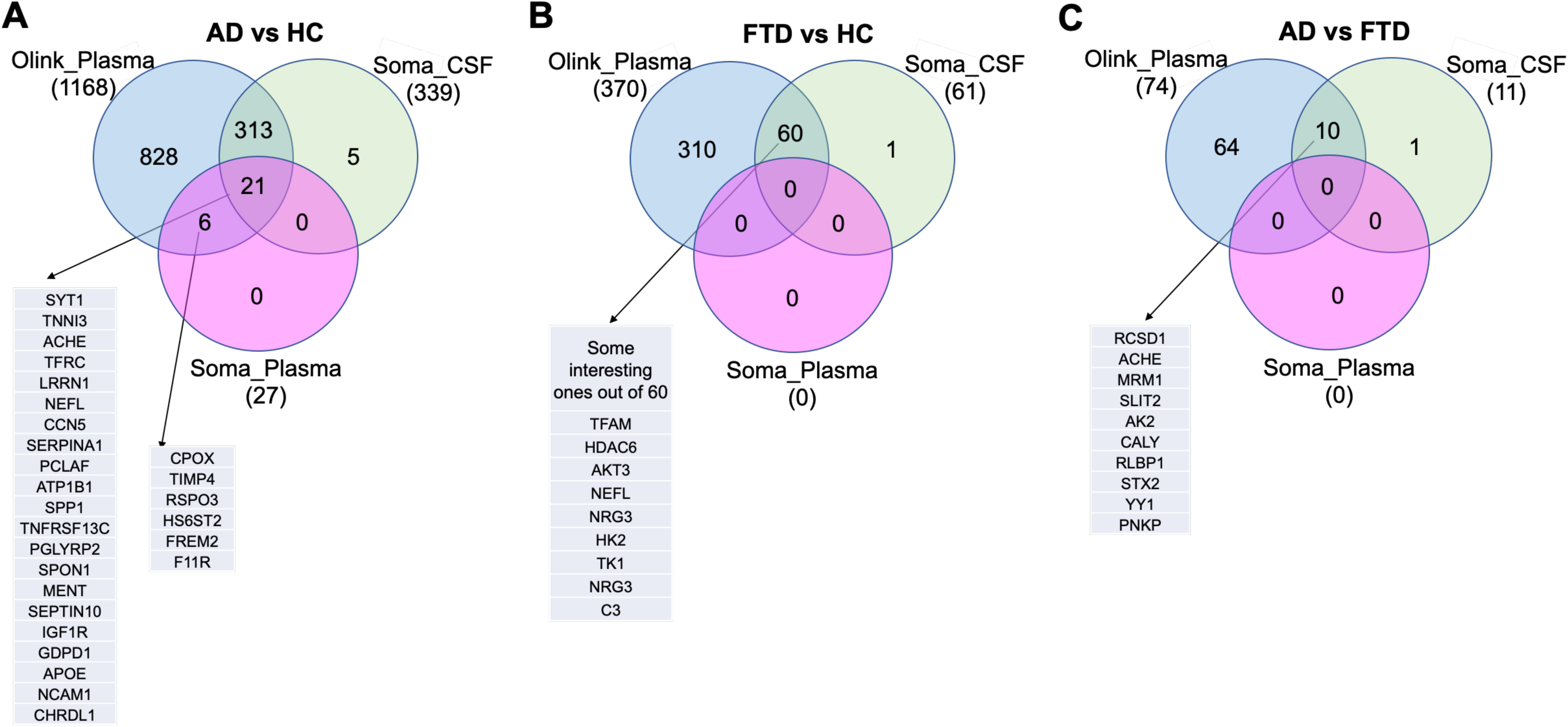
Cross-platform validation of differentially abundant plasma proteins in AD and FTD. Venn diagrams showing the overlap of DAPs identified in the present Olink Explore HT dataset with proteins detected in independent SomaScan plasma and CSF cohorts. Each panel represents a disease comparison: (A) AD vs HC, (B) FTD vs HC, and (C) AD vs FTD. Blue circles denote DAPs identified in our Olink dataset, green circles represent proteins detected in the SomaScan CSF validation cohort, and pink circles indicate the proteins detected in SomaScan plasma cohort. Numbers within each section indicate the count of proteins unique or shared between platforms. Arrows presenting the overlapping proteins (detailed list in Supplementary Table S9).

For FTD, plasma-CSF datasets included 141 unique proteins (174 analytes) of which 67 analytes were FDR-significant. In FTD vs HC comparison 60 proteins were shared between Olink plasma and SomaScan CSF, but no proteins were common across all three datasets highlighting greater molecular divergence and reduced cross-cohort stability in FTD relative to AD, likely due to the limited sample size. The shared plasma–CSF proteins included NEFL, TFAM, HDAC6, AKT3, FGF16, FGFR2, C3 and NRG3 implicating axonal injury, mitochondrial dysfunction, cytoskeleton regulation and neuro-immune signaling as reproducible proteins detectable across biofluids.

Direct AD-FTD plasma-CSF analysis, out of 28 unique proteins only 11 SomaScan analytes passed FDR correction, while no proteins reached FDR significance in the plasma–plasma analysis. When FDR passed SomaScan analytes were intersected with olink plasma data, only 10 common proteins were found including ACHE, SLIT2, CALY and YY1. This again, underscores the challenge of identifying robust cross-platform plasma biomarkers for differential dementia diagnosis.

#### 3.6.3 Comparison with other published proteomic studies

To further assess reproducibility in an ethnic Chinese population, we compared our findings with Olink 1600 plasma proteomic study conducted in Hong Kong Chinese cohort^5^. Among the top proteins identified in our study, PPP1R9B (also known as Spinophilin), a dendritic spine protein regulating synaptic plasticity, was found to be downregulated in both studies^5^. The finding is particularly significant given the established role of synaptic dysfunction and loss in the progression of AD and other neurodegenerative diseases. Notably, PPP1R9B was also downregulated in FTD in our cohort, supporting synaptic degeneration as a convergent mechanism across dementias.

Another overlapped protein is PLXNA4 (Plexin-A4), a receptor for class 3 semaphorins, regulating axon guidance, dendritic spine remodelling, and synaptic pruning. This protein is not significantly dysregulated in FTD in our cohort, supporting disease specificity. Other overlapping proteins from top-ranked AD-associated proteins in Hong Kong study that were significant in our cohort included CD69, SNAP29, BCR1, BANK1, INPPL1, CLIP2 (downregulated) and PAPPA and PTX3 (upregulated). DKK4 showed an opposite direction of effect. We further compared our results with the 19 plasma hub proteins proposed in that study to differentiate AD from the HC. Among those, NELL1 and CD164 were consistently downregulated in both cohorts while GAMT and NFKBIE showed opposite regulation in our data (Supplementary Figure 3 A, B).

Furthermore, a recent study using the Olink 5k platform in CSF applied pseudotime modeling and identified several proteins, including SDC4, MAPT, SMOC1, and ITGB2, as key predictors of disease progression^50^. Among these, we also observed significant alterations in plasma MAPT, DTX3, ITGAM, ACHE, and CEND1 in AD, suggesting partial overlap between CSF- and plasma-derived signals despite differences in biofluid and analytical approaches. Importantly, because these proteins were closely linked to predicted trajectories of future disease progression in CSF, their dysregulation in plasma strengthens their potential utility as accessible markers of AD progression.

## 4. DISCUSSION

Using high-throughput and high-precision immunoassay technique, we systematically profiled over 5400 plasma-proteins to investigate the biofluid proteomic landscape of clinically and biomarker confirmed AD and FTD^13^. Our findings successfully replicated several known AD-associated plasma proteins, including GFAP, BACE1, and ACHE while identify hundreds of previously unreported altered proteins in AD. This study uniquely identifies novel protein alterations in FTD, an area with a limited number of previous proteomics studies^6,7^, thereby contributing new insights to the understanding of both diseases. Integration with single-cell transcriptomic datasets further demonstrated that these plasma signatures reflect contributions from multiple brain cell types-including neuronal, glial, and vascular populations-highlighting the multifactorial cellular basis of both diseases. Together these features make our dataset a valuable contribution towards mechanistic understanding and global biomarker generalizability.

Major strength of our AD findings is the convergence of differential expression, machine learning, and cell-type integration on biological process that are important in AD pathogenesis. Among the most significantly upregulated proteins were GFAP, BACE1, ACHE, HDAC6, PPP1R9B, BENC2 and SYT1, each of which has established links to neurodegeneration. GFAP, a marker of astrocytic activation, is consistently elevated in the CSF and plasma of individuals with AD and tracks amyloid-β pathology and neurodegeneration^51^. In our cohort, the significant increase in plasma GFAP and its strong correlation with p-tau species and cognitive performance further supports the link between reactive gliosis and tau-driven neurodegeneration and clinical decline^52^. Similarly, BACE1, a key enzyme initiating amyloid-β peptide generation was also elevated with its significant correlation Aβ_42_ levels, supporting its key role in amyloidogenic processing.

Beyond these established biomarkers, we found several mechanistically important proteins in our data. HDAC6, a cytoplasmic deacetylase regulating microtubule stability and autophagy, was upregulated in AD brain tissue and contributes to tau hyperphosphorylation and impaired axonal transport, and its increase in AD plasma, aligns with its reported overexpression in AD. PPP1R9B (spinophilin), a postsynaptic scaffolding protein critical for dendritic spine structure and synaptic plasticity was reduced in AD plasma, consistent with synapse loss and cognitive decline. Similarly, reduced BECN1 (Beclin-1) levels support impaired autophagic clearance pathways^25^ while increased SYT1 reflects presynaptic dysfunction previously observed in CSF studies^26^. Collectively, this indicates that plasma proteomics captures multiple pathologies in AD including gliosis, amyloid processing, synaptic degeneration and disrupted proteostasis.

Multivariate GLMNET analysis delineates distinct diagnostic molecular architectures for AD and FTD. Classification of AD versus healthy controls was driven primarily by markers of astrogliosis (GFAP)^21,51^ and amyloid-related processes (BACE1, ACHE)^22,45^. Interestingly, many of these proteins were independently replicated in large scale external cohorts. Cross platform validation with GNPC SomaScan dataset^7^ showed strong concordance across 232 proteins including metabolic stress, genomic instability, and neuroinflammation (HK2^53,54^, DHX8^55^, TNF^56^), alongside indicators of endothelial dysfunction, blood–brain barrier (BBB) dysfunction, and broader vascular compromise (PECAM1^57^, ENG^58^). Some of these proteins also have known AD associations: SPON1 affects APP processing and has been linked to slower cognitive decline^59^, while NCAM1 supports synaptic adhesion and plasticity and is disrupted in AD^60^. The high stability of these reproducible markers suggests they reflect fundamental systemic features of AD pathophysiology, making them prime candidates for both biomarkers and therapeutic targets.

When comparing the Olink-5k plasma and SomaScan-7k CSF/plasma datasets, several overlapping proteins have previously been associated with AD (SYT1, SPP1, APOE, ACHE, NEFL, TFRC, and IGF1R), supporting the biological relevance of our findings. Many others remain poorly characterized in this context and therefore represent promising novel biomarker candidates. Notably, some proteins from the overlap analysis (CCN5, LRRN1, and NCAM1) have emerging links to neural structure, vascular remodeling, and synaptic connectivity^60–62^. Synaptic cell-adhesion molecules such as NCAM1 are disrupted in AD^60^. Similarly, matricellular regulators like CCN5 promote neurite outgrowth and may implicate synaptic connectivity pathways in neurodegeneration^62^. LRRN1, a neuronal leucine-rich repeat protein involved in neurite outgrowth and synaptic connectivity, has recently been identified in multiple large-scale AD blood proteomic studies^7,19^. Notably, partial concordance with CSF pseudotime-modeling studies^50^ underscores that plasma can also reflect proteins associated with disease progression trajectories including SDC4, MAPT, SMOC1, and ITGB2, as highly important predictors of disease progression^50^. The high effect-size correlation across platforms supports the stability and generalizability of the AD plasma proteome and indicates that these signals are not assay-specific.

As compared to AD, FTD has not been studied as much in large scale plasma proteomics. Our analysis revealed a more stringent but biologically distinctive signature for FTD. The FTD profile was defined by complement-mediated inflammation (C3)^32^ and overt axonal degeneration (NEFL)^33^. Direct discrimination between AD and FTD required a more specific molecular panel beyond these general markers. While GFAP and ACHE remained discriminators, the AD–FTD model highlighted additional features such as PCYT1A (lipid biosynthesis)^63^ and SLIT2 (axon-guidance)^64,65^. These findings indicate that accurate differential diagnosis between dementia subtypes depends on capturing disease-specific disruptions in lipid metabolism and synaptic/axon wiring, rather than relying solely on shared markers of neuronal injury. Our data also identified proteins implicating mitochondrial dysfunction (TFAM, AKT3), apoptosis (BCL2L11), inflammatory lipid metabolism (LTA4H) and synaptic/ neurotrophic pathways (PPFIA2, NTF4).

Pathway enrichment of FTD proteins highlighted nuclear and DNA metabolic processes, innate immune activation, and aberrant RAS–RAF–MAPK and PI3K/AKT signaling biological themes aligning with genetic and pathological studies of FTD implicating RNA/DNA processing, protein homeostasis, and dysregulated kinase signaling^66^. The enrichment of FGFR2-mediated pathways and ubiquitin-specific proteases further supports the relevance of growth factor signaling^67^ and proteostasis dysfunction^68^. When compared with other datasets^7^ our FTD data also showed strong concordance. Concordant proteins were also involved in FGFR2/3/4 and IGF1R signaling, ubiquitin-processing pathways, DNA damage repair, and transition metal ion binding. Importantly, our study expands the FTD biomarker landscape beyond NEFL by introducing novel candidate proteins that may support mechanistic stratification and therapeutic development.

Comparative pathway analyses revealed both convergent and disease-specific molecular alterations across AD and FTD. While AD-unique proteins were enriched for mitochondrial dysfunction, cytoskeletal organization, and synaptic processes, and FTD-unique proteins highlighted DNA repair, innate immune signaling, and growth factor–kinase pathways, the overlapping proteins pointed to a shared core of cellular dysregulation. The consistent enrichment of phosphatidylinositol metabolism, ER–Golgi trafficking, and ubiquitin-processing proteases suggests that perturbations in membrane lipid signaling, intracellular transport, and proteostasis represent common molecular denominators in both diseases. These shared mechanisms likely converge on synaptic dysfunction and neuronal vulnerability, providing a mechanistic bridge between the distinct pathological signatures of AD and FTD and highlighting potential cross-disease therapeutic targets that stabilize vesicular trafficking and lipid-signaling homeostasis. PPP1R9B (spinophilin) particularly provides an example of synaptic degeneration as convergent pathway in both diseases. Its decrease in AD^69,70^ and FTD supports synaptic degeneration as a shared endpoint across neurodegenerative dementias.

This study represents one of the first large-scale plasma proteomic analyses of AD and FTD in a Southeast Asian cohort. Dementia proteomics research has been conducted mostly in populations of European ancestry, leaving Asian populations cohorts substantially underrepresented. Existing Asian plasma proteomics studies in AD have been limited and have used smaller protein panels^5^. We observed overlaps of several synaptic and inflammatory proteins, but limited size and scope of previous studies preclude firm conclusions regarding ethnic-specific differences. Moreover, our top AD markers largely coincide with those reported in predominantly Caucasian cohorts, supporting cross-ethnic generalizability of core disease biology. Our findings also extend prior plasma proteomic work on predictive signatures for cognitive decline in Southeast Asian cohorts^71^ by using pTau217 and pTau218 confirmed AD and FTD cohort. Together, these data represent an important step towards global coverage in biomarker research and provide a reference resource for future comparative studies.

One major limitation is the modest sample size, particularly in FTD. Given the small sample sizes, further validation using more patient data would be needed. However, with the high cost and technical demands of large-scale Olink HT-Explore profiling required in smaller discovery cohorts, we tried to compensate this by stringent biomarker characterization and extensive external validation. Some degree of model overfitting is expected; nevertheless, the coefficient plots remain informative and are broadly concordant with existing literature. In addition, different proteomic platforms employ distinct detection chemistries like Somascan uses multiple aptamers targeting per protein^72^. The unique protein overlaps with our Olink data was defined only based on FDR-significant Olink targets, with corresponding Somascan analytes evaluated where available. Differences in overlap between plasma and CSF datasets likely reflect biofluid-specific quality control and panel composition. Future longitudinal plasma proteomics studies will be essential to assess disease progression and predictive utility. In summary, comprehensive plasma proteomics profiling of biomarker-confirmed AD and FTD identified distinct and shared molecular pathways reflecting mitochondrial dysfunction, synaptic degeneration, immune activation, lipid signaling disruption and proteostasis deregulation. Validation across independent datasets supports the robust and assay-independent plasma signature of AD, while the identification of novel FTD-associated proteins expands the currently limited biomarker landscape of this disorder. Reproducibility across platforms and populations indicates that plasma proteomic signatures may serve as scalable biomarkers suitable for large-scale screening, longitudinal monitoring, and therapeutic trial stratification. These findings provide a foundation for scalable blood-based differential diagnostics and for targeting shared and disease specific molecular features in neurodegenerative dementias.

## Data Availability

All data produced in the present study are available upon reasonable request to the corresponding author.

## Acknowledgements

The authors sincerely thank all participants and research staff, especially Shermine HL Low, Louis SS Lim, and Valerie JZ Teh from the Department of Research, National Neuroscience Institute, Singapore, for their invaluable contribution to this study. We also extend our appreciation to Regina Chen and Yi Hao Nah from the Department of Research, National Neuroscience Institute, Singapore, for their efforts in recruiting healthy controls utilized in our research.

## Consent statement

The written informed consent was obtained from each participant prior to recruitment into the study. This study was approved by the SingHealth Institutional Ethics Review Board and certify that the study was performed in accordance with the ethical standards as laid down in the 1964 Declaration of Helsinki and its later amendments or comparable ethical standards.

