## Supplemental Figures for "Deep Plasma Proteomics Reveals Shared and Disease-Specific Molecular Signatures in Alzheimer’s Disease and Frontotemporal Dementia"

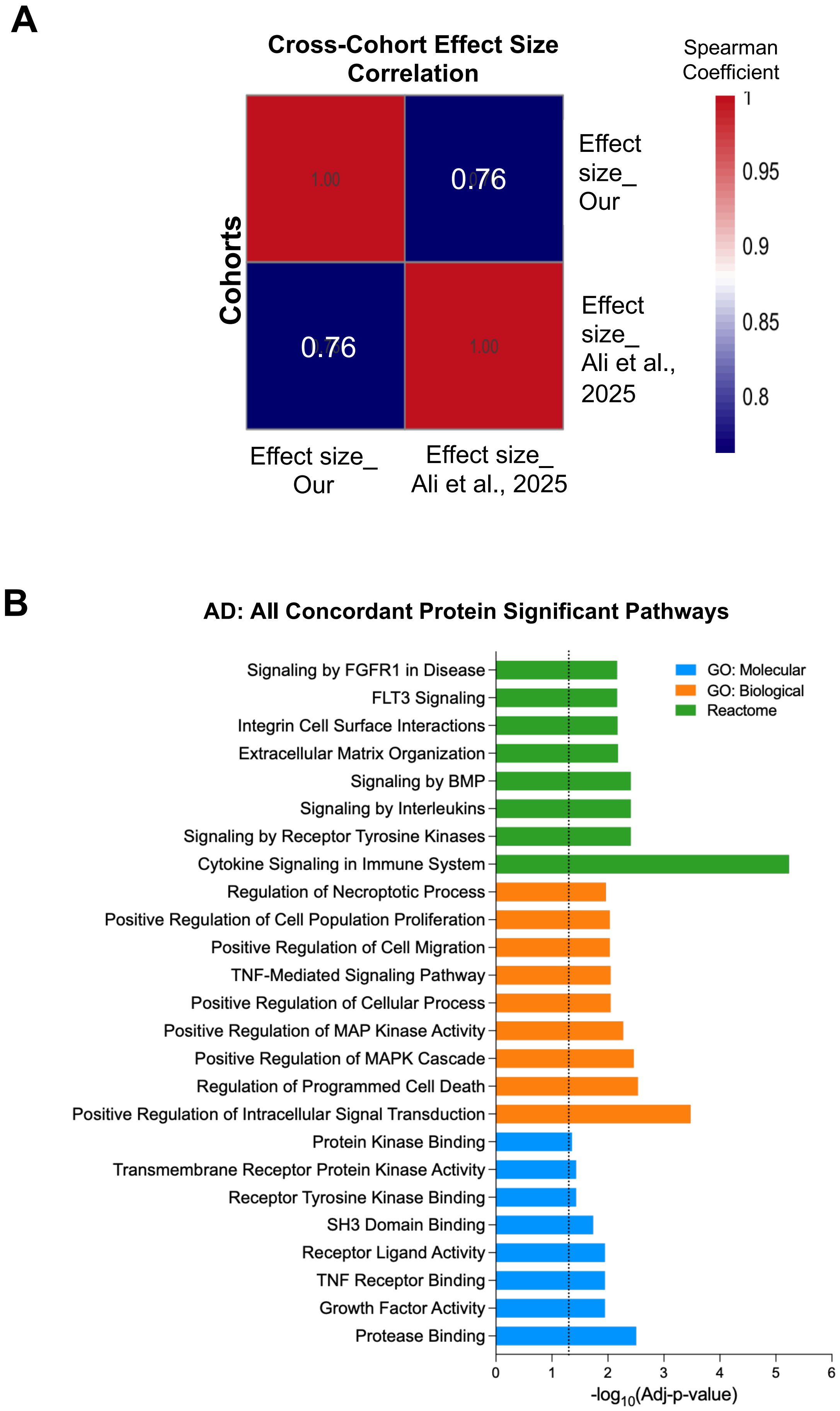


**Supplementary Figure 1.** Cross-cohort validation of DAPs in AD vs HC comparison. **A.** Heatmap summarizing cross-cohort effect size correlation. Spearman correlation analysis shows high similarity between studies (ρ=0.76). **B.** Pathway enrichment for the concordant proteins showing significantly enriched GO molecular functions, biological processes and reactome pathways.

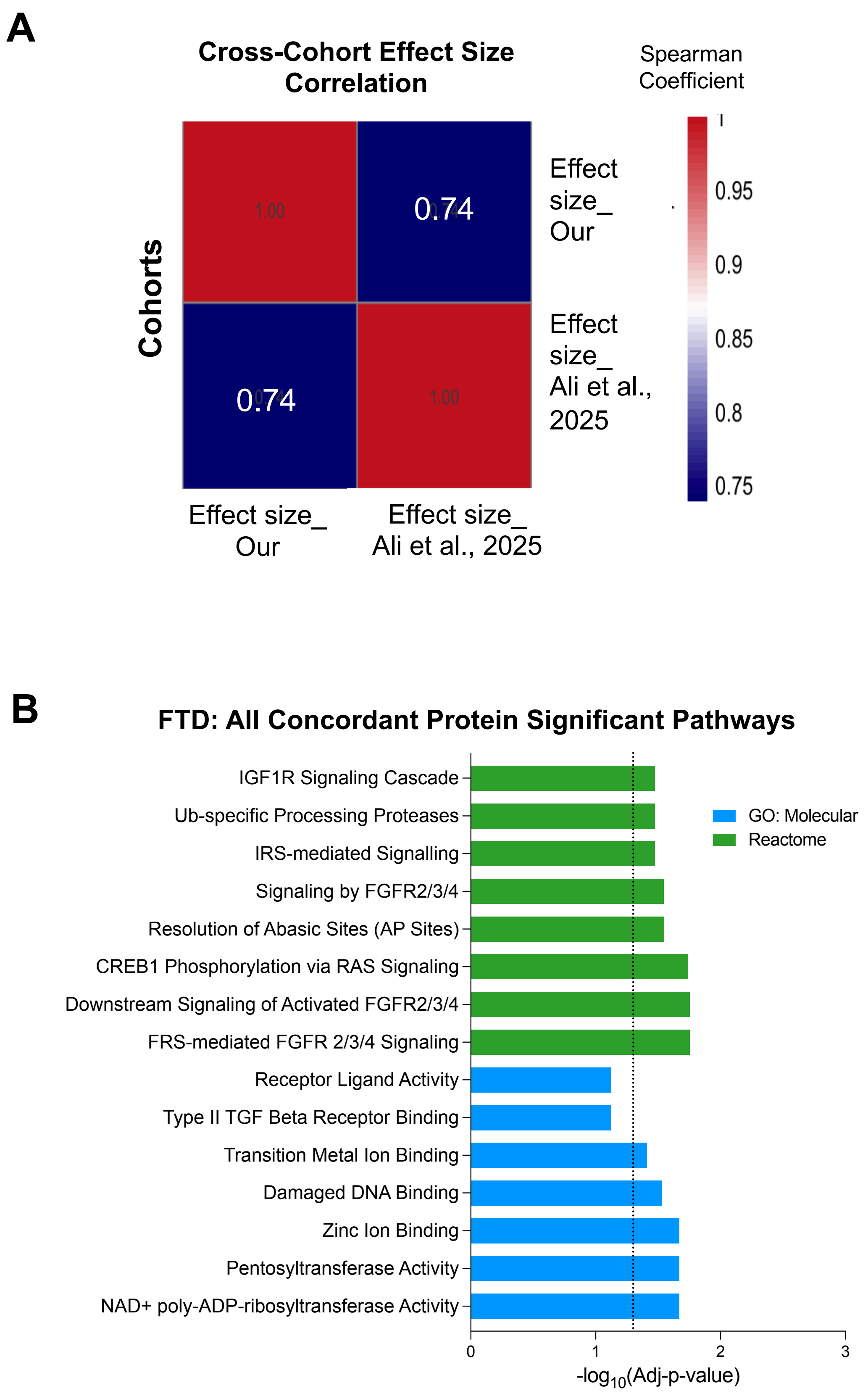


**Supplementary Figure 2.** Cross-cohort validation of DAPs in FTD vs HC comparison. **A.** Heatmap summarizing cross-cohort effect size correlation. Spearman correlation analysis shows high similarity between studies (ρ=0.74). **B.** Pathway enrichment for the concordant proteins showing significantly enriched GO molecular functions and reactome pathways.

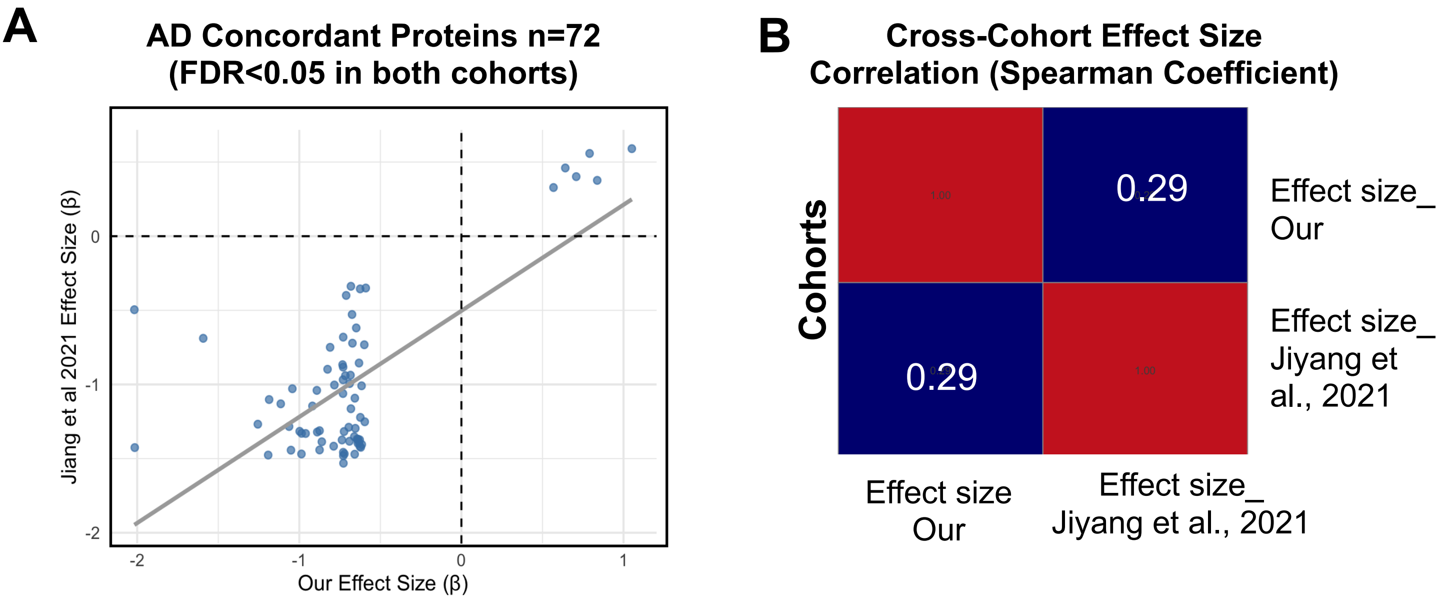


**Supplementary Figure 3.** Cross-cohort validation of DAPs. FDR <0.05 selected DAPs in AD vs HC were compared with external published data. **(A)** Scatter plot comparing $\log_2$ fold-change effect sizes (β) from the present study ("Our") with those reported in an independent external Hong Kong cohort. Each point represents a shared protein between platforms. Gray regression line indicates the linear relationship between studies. Dashed lines denote zero effect in each dataset. A positive association demonstrates concordant direction and magnitude of protein alterations across cohorts. **(B)** Heatmap summarizing cross-cohort effect size correlation. Spearman correlation analysis shows slight similarity between studies (ρ=0.29).
